# Learning from the resilience of hospitals and their staff to the COVID-19 pandemic: a scoping review

**DOI:** 10.1101/2021.04.22.21255908

**Authors:** Lola Traverson, Jack Stennett, Isadora Mathevet, Amanda Correia Paes Zacarias, Karla Paz de Sousa, Andrea Andrade, Kate Zinszer, Valéry Ridde

## Abstract

**Background:** The COVID-19 pandemic has brought huge strain on hospitals worldwide. It is crucial that we gain a deeper understanding of hospital resilience in this unprecedented moment. This paper aims to report the key strategies and recommendations in terms of hospitals and professionals’ resilience to the COVID-19 pandemic, as well as the quality and limitations of research in this field at present.

**Methods:** We conducted a scoping review of evidence on the resilience of hospitals and their staff during the COVID-19 crisis in the first half of 2020. The Stephen B. Thacker CDC Library website was used to identify papers meeting the eligibility criteria, from which we selected 65 publications. After having extracted data, we presented the results synthesis using an “effects-strategies-impacts” resilience framework.

**Results:** We found a wealth of research rapidly produced in the first half of 2020, describing different strategies used to improve hospitals’ resilience, particularly in terms of 1) planning, management, and security, and 2) human resources. Research focuses mainly on interventions related to healthcare workers’ well-being and mental health, protection protocols, space reorganization, personal protective equipment and resources management, work organization, training, e-health and the use of technologies. Hospital financing, information and communication, and governance were less represented in the literature.

**Conclusion:** The selected literature was dominated by quantitative descriptive case studies, sometimes lacking consideration of methodological limitations. The review revealed a lack of holistic research attempting to unite the topics within a resilience framework. Research on hospitals resilience would benefit from a greater range of analysis to draw more nuanced and contextualized lessons from the multiple specific responses to the crisis. We identified key strategies on how hospitals maintained their resilience when confronted with the COVID-19 pandemic and a range of recommendations for practice.

## INTRODUCTION

Since the end of 2019, the emergence and rapid spread of COVID-19 has put health systems across the globe under severe strain [1]. The COVID-19 is rapidly evolving, and its many unknowns have made the response efforts difficult and variable, challenging effective decision-making in a context of uncertainty [2], [3]. National responses have varied greatly, with some countries being more successful (i.e., better prepared, or better able to cope with the pandemic) than others in containing the transmission and preventing deaths. In other words, some health systems have been more *resilient* to the shock of the pandemic [4].

Hospitals play a critical role within health systems in providing essential medical care to the community, particularly during a crisis [5]. They are complex and vulnerable institutions, dependent on crucial external support and supply lines [6]. Many of them frequently operate at near-surge capacity under normal working conditions, and a rise in admission volume, even modest, can overwhelm them beyond their functional reserve [5], [7]. The current outbreak of COVID-19 has caused numerous hospitals’ disruptions such as shortages of critical equipment and supplies [8], [9], significant risks of infection among patients or healthcare workers (HCWs) [10], [11], high hospitalization rates and lengths of stay [12], and high workloads for HCWs that are often linked to psychological adverse effects [13], [14]. All those disruptions can have a direct impact on healthcare delivery by limiting access to needed care, creating panic and potentially jeopardizing established working routines [5].

Understanding health systems resilience has never been more essential than today [15]. Resilience refers to “the capacities of dimensions/components of a health system faced with shocks, challenges/stress or destabilizing chronic tensions (unexpected or expected, sudden or insidious, internal or external to the system), to absorb, adapt and/or transform in order to maintain and/or improve access (for all) to comprehensive, relevant and quality health care and services without pushing patients into poverty” [16], [17]. To our knowledge, despite a growing interest in the concept of resilience, little work has explored how it has been operationalized in empirical studies [18]. Except for a few studies focusing on HCWs’ mental health and their personal resilience, there has been no attempt to systematically summarize the evidence on improving and maintaining hospitals resilience generated from the COVID-19 outbreak.

Moreover, in a context of a public health emergency, research and action co-evolve and are conducted at the same time. In the context of COVID-19, the scale of scientific production has been astonishing and, while essential, it can be overwhelming and counterproductive [19]. It is necessary to find effective ways to share methodological sound information between countries and stakeholders during this crisis management.

This paper aims to report the main strategies implemented by hospitals and their staff to face the COVID-19 pandemic worldwide. It is not intended to be exhaustive, but its overall purpose is to offer evidence on best practices and lessons learned from hospitals’ experiences to guide health decision makers and professionals to design adapted and efficient responses to cope with similar shocks in the future [20].

## METHODS

As part of a multi-country analysis [21], we conducted a scoping review, based on the highly empirical scientific literature published in the first half of 2020. A scoping review was preferred to a full systematic review as it allowed us to synthesize, from a very broad search, with rigor and in a relatively short period of time, the state of knowledge on a specific research question and to identify and analyze gaps in the knowledge base to inform public decision makers, stakeholders, and researchers [22], [23].

The methods of data selection and analysis have been detailed in an online protocol [24]. Although not always applicable, we referred to the Preferred Reporting Items for Systematic reviews and Meta-Analyses extension for Scoping Reviews (PRISMA-ScR) checklist [25].

### Identification of articles

We conducted our searches on a collection of articles related to the COVID-19 pandemic published on the Stephen B. Thacker CDC Library website. These articles were collected on the following electronic databases : Medline (Ovid and PubMed), PubMed Central, Embase, CAB Abstracts, Global Health, PsycInfo, Cochrane Library, Scopus, Academic Search Complete, Africa Wide Information, CINAHL, ProQuest Central, SciFinder, the Virtual Health Library, LitCovid, WHO COVID-19 website, CDC COVID-19 website, China CDC Weekly, Eurosurveillance, Homeland Security Digital Library, ClinicalTrials.gov, bioRxiv (preprints), medRxiv (preprints), chemRxiv (preprints), and SSRN (preprints). We downloaded, from April to June 2020, all the references added to the CDC database. The first references added to the database were published in December 2019.

### Selection of studies

After downloading downloaded all the references from the CDC database (n = 58160), we imported them on Zotero, a reference management software, to make a first sorting with an English request (Table 1). Requests in French and Spanish proved not to be relevant or effective.

**Table 1:**
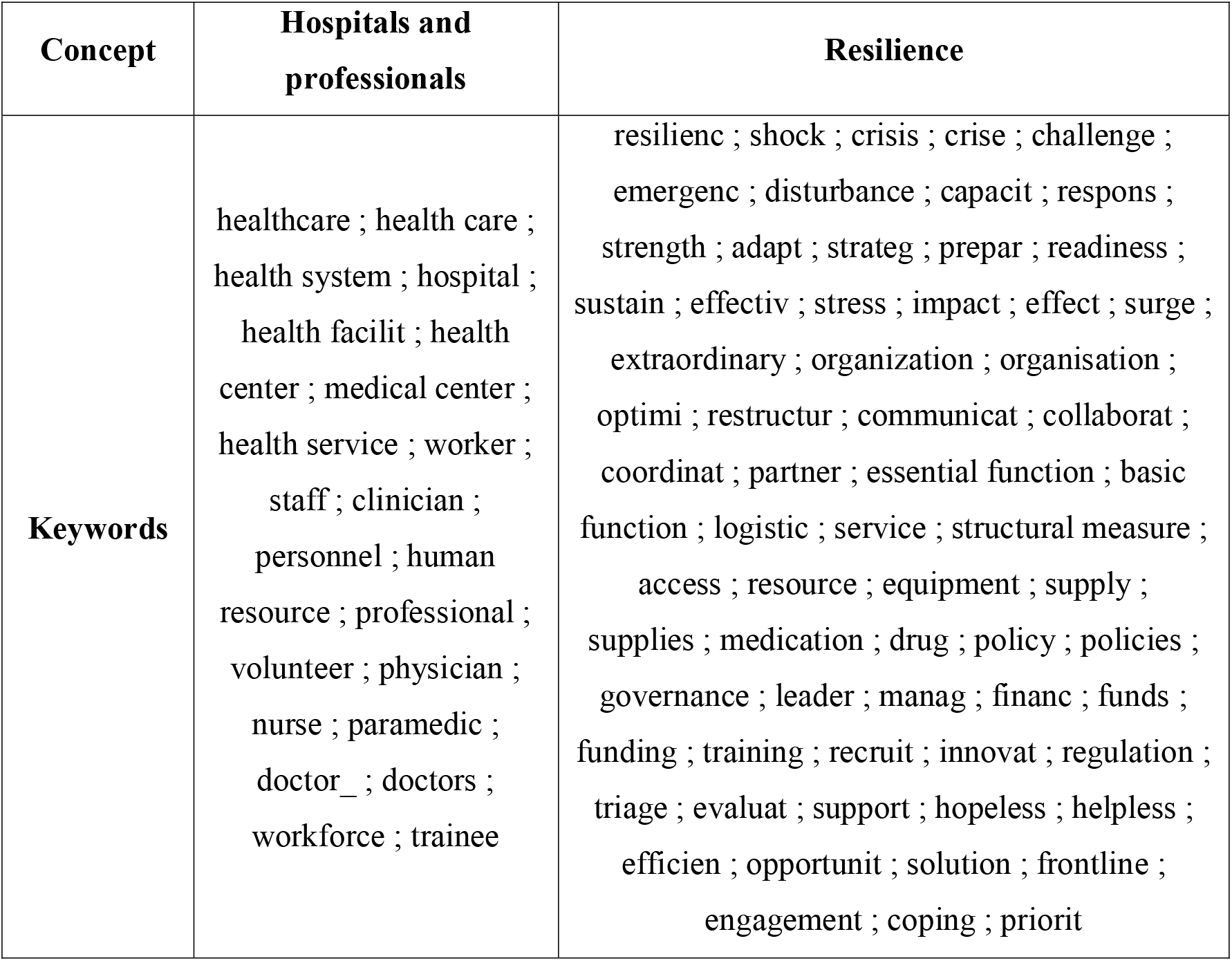
English request on Zotero.

| Concept | Hospitals and professionals | Resilience |
| --- | --- | --- |
| **Keywords** | healthcare ; health care ;<br>health system ; hospital ;<br>health facilit ; health<br>center ; medical center ;<br>health service ; worker ;<br>staff ; clinician ;<br>personnel ; human<br>resource ; professional ;<br>volunteer ; physician ;<br>nurse ; paramedic ;<br>doctor_ ; doctors ;<br>workforce ; trainee | resilienc ; shock ; crisis ; crise ; challenge ;<br>emergenc ; disturbance ; capacit ; respons ;<br>strength ; adapt ; strateg ; prepar ; readiness ;<br>sustain ; effectiv ; stress ; impact ; effect ; surge ;<br>extraordinary ; organization ; organisation ;<br>optimi ; restructur ; communicat ; collaborat ;<br>coordinat ; partner ; essential function ; basic<br>function ; logistic ; service ; structural measure ;<br>access ; resource ; equipment ; supply ;<br>supplies ; medication ; drug ; policy ; policies ;<br>governance ; leader ; manag ; financ ; funds ;<br>funding ; training ; recruit ; innovat ; regulation ;<br>triage ; evaluat ; support ; hopeless ; helpless ;<br>efficien ; opportunit ; solution ; frontline ;<br>engagement ; coping ; priorit |

All the references selected on Zotero (n = 13173) have been classified by the Automated Text Classifier of Empirical Research (ATCER) tool. The ATCER tool distinguishes (a) empirical studies (with an empirical degree ≥ 50), based on qualitative, quantitative, and mixed methods, from (b) non-empirical work (with an empirical degree < 50) (e.g., commentaries, editorials, literature reviews, professional guidelines etc.) [26]. Because of the large number of data, we selected an ATCER threshold of 90 to reflect articles that were judged as “highly empirical” by the tool (i.e., with an empirical degree ≥ 90). Grey literature and pre-publications were excluded from our research.

To be included in the review, the references had to meet the following criteria: (a) published between December 2019 (the beginning of the COVID-19 pandemic) and June 2020; (b) written in English; (c) assessed as “highly empirical” by the ATCER tool (i.e., with an empirical degree ≥ 90); (d) available and accessible in full-text; (e) focused on the resilience of hospitals and their staff to the COVID-19 pandemic.

After the ATCER tool analysis and the removal of duplicates, we imported the selected references (n = 559) into Covidence, a systematic review software for screening. Covidence automatically removed 1 duplicate. Two reviewers (LT, IM) independently assessed the relevance of titles and abstracts based on the inclusion and exclusion criteria. In case of disagreement between the two reviewers, a third reviewer (JS) made a final decision. At this stage, 411 references were excluded. Reasons for exclusion are detailed in the PRISMA flow diagram (Figure 1). Then, at least two of five involved reviewers (LT, JS, AC, KP, AA) independently assessed the full text relevance of the selected articles. After a common vote, they agreed to exclude 82 references. We finally included 65 references in the review.

**Figure 1:**
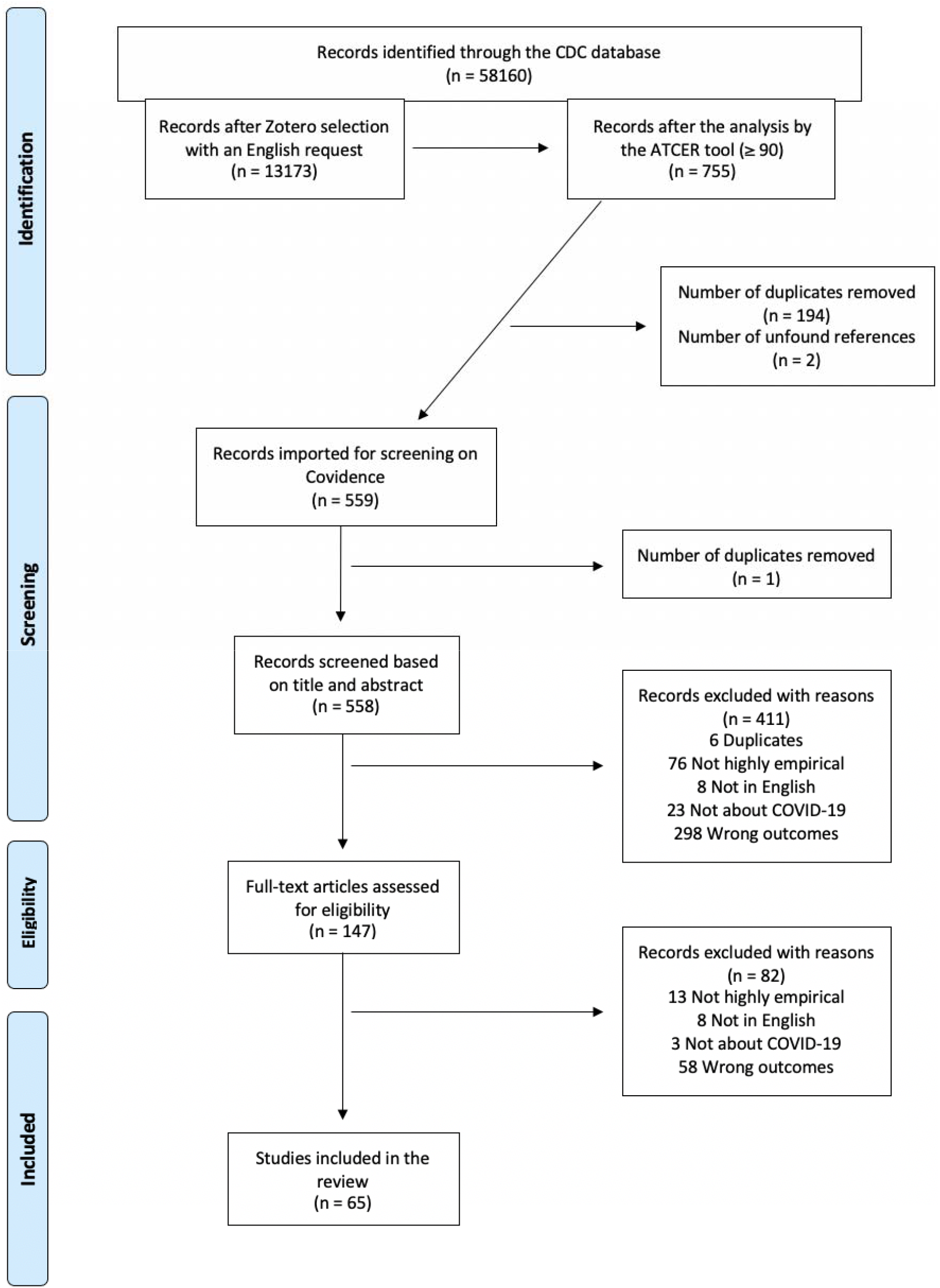
PRISMA flow diagram.

### Data extraction and quality assessment

One author (LT) extracted data from the selected studies (n = 65) on MAXQDA® 2020, a software package for qualitative, quantitative and mixed methods research.

A data extracting form was created on Excel to collect several information from the selected literature: publication type, study type, study settings – continent and hospital settings –, hospital dimension(s), objectives, results and limitations of the study, and conceptual framework or mid-range theory used (Table S1: Description of the selected studies).

Information related to quality assessment of the studies with the Mixed-Methods Appraisal Tool (MMAT) were reported in (Figure 3).

### Data synthesis and analysis: the conceptual approach

To synthetize data and write the review, we used a conceptual framework on health systems’ resilience designed by Ridde and al. [21].

The framework shows the association between: (a) the effects, positive or negative, caused by the pandemic; (b) the strategies implemented to deal with these effects; and (c) the impacts, positive or negative, of these strategies on the hospital’s organizational routines.

From a resilience perspective, these “Effects – Strategies – Impacts” processes can give rise to three types of configuration that could be named as: (a) reaction (i.e., an effect is felt, a strategy is adopted, and this strategy has an impact), (b) anticipation (i.e., strategies/impacts, before any effects), and (c) inaction (i.e., effects but no strategies). These processes are nonlinear and the impact of one strategy can, for example, lead to the implementation of a new strategy. We do not make any judgements about the nature of the causality of the impacts and the evaluations that provide evidence for them. Thus, we only describe the impacts of the strategies as proposed by the authors, and we will report any methodological limitations in the Discussion section.

We adapted the 10 dimensions of health systems resilience [27] to analyze hospitals and professionals’ resilience to the COVID-19 pandemic, and we chose to focus on the 5 following dimensions: 1) governance; 2) human resources; 3) finance; 4) planning, management, and security; 5) information and communication.

## RESULTS

We selected 65 articles for inclusion in the review (Figure 1). We mapped the distribution of the selected studies by publication type, study type, continent/country of the study, hospital settings, hospital dimension(s) and the use of conceptual frameworks or mid-range theories (Table S1: Description of the selected studies).

Our analysis revealed that 95.4% (n = 62) of the articles were identified as peer-reviewed articles. The three other studies were peer-reviewed short or case reports. The studies were dominated by descriptive quantitative studies (n = 40; 61.5%) and case studies, with single case studies (n = 18; 27.7%) and multiple-case studies (n = 5; 7.7%) using both quantitative (n = 19; 29.2%) or mixed methods (n = 4; 6.1%). There were only 1 randomized study (1.5%), and 1 qualitative study (1.5%).

The geographical distribution of the selected studies is described in (Figure 2). The studies were conducted in Asia (n = 35; 53.8%) with most studies conducted in China (n = 22; 33.8%), in Europe (n = 12; 18.5%) with most studies conducted in Italy (n = 4; 6.1%) and Spain (n = 3; 4.6%), in North America (n = 8; 12.3%), in Latin America (n = 2; 3%), and in Oceania (n = 2; 3%). The remaining studies (n = 5; 7.7%) were conducted in multiple countries. Location was not mentioned for one study.

**Figure 2:**
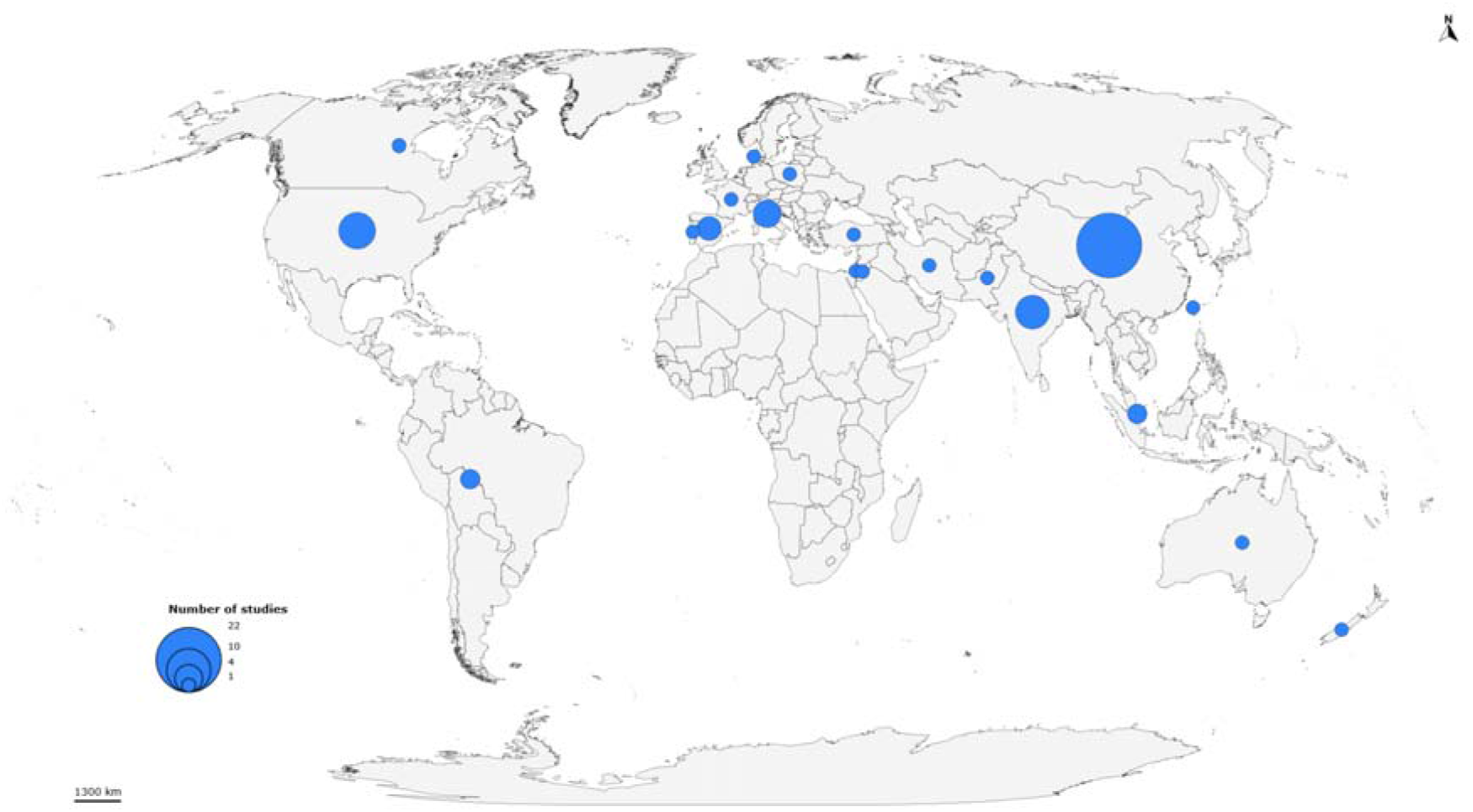
Geographic distribution of identified studies.

Most of the studies focused on hospital settings (n = 33; 50.8%), with 12 studies focusing on tertiary hospitals (18.5%), or on both public and private practice settings (n = 12; 18.5%). Several studies focused on specific departments or units (n = 27; 41.5%), and mostly on emergency or infectious departments (n = 13; 20%).

In the selected studies, hospital dimensions were referred to at the following frequency (some studies focused on more than one dimension): 1) planning, management, and security (n = 77), with most studies focusing on protection protocols (n = 24); 2) human resources (n = 58) with most studies focusing on professionals’ well-being and mental health (n = 32); 3) information and communication (n = 7); and 4) finance (n = 4). No article specifically focused on the “governance” dimension (i.e., on hospitals’ leadership decisions) but we found some elements concerning governance in studies that focused on the 4 other dimensions.

We applied the MMAT to report quality assessment of the studies (Figure 3). Almost all studies had clear described research questions or objectives. Sampling methods or techniques were clearly mentioned in 8 studies (12.3%). Representation of the population under study seems to have been well addressed in most of the studies, but we found a high representation of female respondents in some studies (n = 20; 30.8%), and a high representation of male respondents in some other studies (n = 7; 10.8%). Appropriate measurement was captured well in almost all the studies, except for 5 of them in which it was not mentioned (7.7%). Response rate was clearly reported in only 9 studies (13.8%) and 52 articles explicitly mentioned limitations of the intervention or of the study methodology (80%).

**Figure 3:**
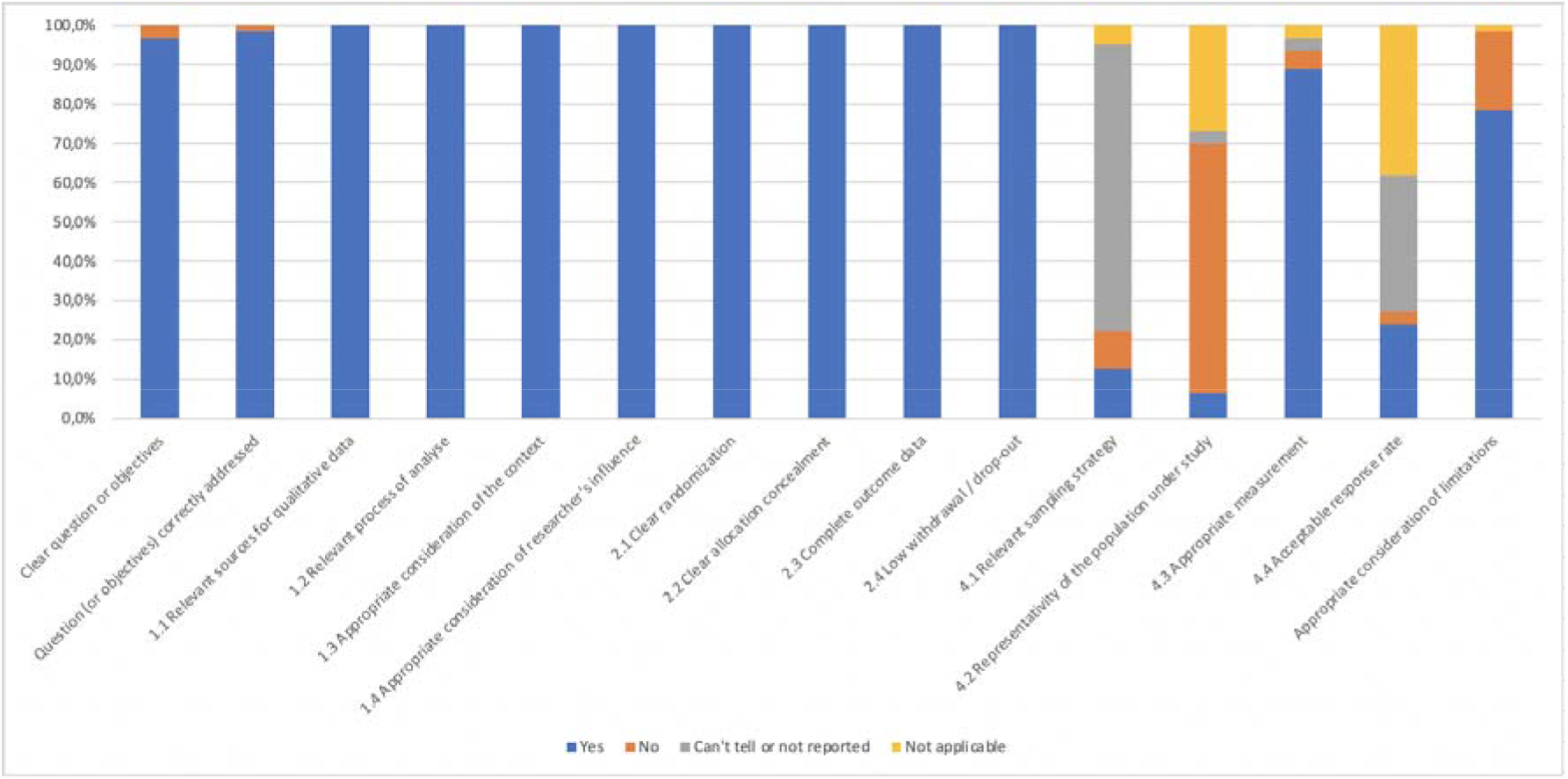
Quality assessment of the selected studies according to the MMAT.

Only 1 study [28] used a conceptual framework to further analyze data.

In the following synthesis, we reported the main “Effects – Strategies – Impacts” processes that appeared in the selected articles to study hospitals’ resilience.

### 1. Hospital dimension: Planning, management, and security

#### 1.1 Protection protocols

##### Effects of the pandemic on hospital organizational routines

The COVID-19 pandemic has urged hospitals to develop and put in place new protocols to preserve the safety of patients and professionals [29]–[32]. Several studies show the high prevalence of infection among healthcare workers (HCWs) that may probably have been infected in their working environment in hospitals [33]. However, safety protocols are not always available in medical institutions receiving COVID-19 patients [9].

##### Strategies implemented to address these effects

- Instituting standard pre-triage and testing protocols. A large tertiary referral center in Northern Italy implemented an out-of-hospital pre-triage to identify suspected SARS-CoV-2 infections [34]. Phone pre-triage was also tested in nuclear medicine (NM) departments for patients with COVID-19 or suspected symptoms (e.g., cough, fever) [35]. The otolaryngology department of the Rush University Medical Center, a tertiary medical center in Chicago, designed and implemented two preoperative COVID-19 testing protocols for all patients undergoing a surgical procedure: a “high risk case protocol” performed two days prior to surgery, and a “universal point-of-car (POC) testing protocol” performed on the day of surgery [36]. Another study led in Australia proposed self-collection of nasal and throat swabs (i.e., done by the patients themselves) as a reliable alternative to HCWs’ collection [37].
- Using algorithms to facilitate patients’ prioritization. In California, a department of surgery of the Stanford University Medical Center created a decision tree algorithm describing institutional guidelines for precautions for operating room team members. The algorithm is based on urgency of operation (disease severity), anticipated viral burden at the surgical site, opportunity for a procedure to aerosolize the virus, and patients’ testing status and symptomatology [38]. The University of Chicago Comer Children’s Hospital implemented a scoring system algorithm for Medically Necessary, Time-Sensitive (MeNTS) procedures, to facilitate case prioritization by incorporating procedure, disease, and patient factors into the decision-making process (e.g., risks of exposing HCWs to potential viral transmission, consumption of scarce medical resources etc.) [39].
- Encouraging HCWs and patients to follow strict hygienic protective measures within hospitals. Professionals and patients were asked to wear adequate personal protective equipment (PPE) (hospital scrubs and footwear for HCWs, and surgical masks for HCWs and patients), and to wash and disinfect their hands [29], [31], [34]. Hospitals were also required to disinfect common work surfaces [29], [40] or to ventilate rooms after each visit [31].
- Avoiding contact between professionals and between HCWs and patients. Some hospitals in Europe and North America suspended, cancelled, or held online clinical sessions and academic committee meetings and conferences [29], [40]. Other hospitals decided to reduce the number of HCWs in rooms hosting suspected and/or confirmed COVID-19 cases [41], or imposed HCWs to complete patients’ medical records once patients have left the room to shorten exposure time [31]. In a study led in Poland, the rule of “one lab, one desk, one telephone” for medical staff was implemented [31].
- Regulating or restricting family visits. Two European hospitals prohibited visits from the relatives of patients [31], [32]. In Taiwan, a large study which analyzed the new visiting policies of 472 hospitals in Taiwan concluded that 276 hospitals posted new visiting policies on their websites. Visits to regular wards were forbidden in 83 hospitals and, among the 193 hospitals that still allowed visits to regular wards, 141 restricted visitors to two at a time and 106 restricted visits to two visiting periods per day [42]. Among these policies, recording patient details regarding recent travel, occupation, and contacts information was mentioned by 159 websites, body temperature monitoring by 151, and hand hygiene by 122.
- Implementing contact tracing of HCWs. In Italy, the University Hospital of Bari implemented a reporting system to record all close contacts of HCWs by a “COVID-19 Control Room” which requests suspected and confirmed HCWs cases to be placed in home isolation and ordains all people who had contact with involved HCWs to do domiciliary nasopharyngeal swabs [41].

##### Impacts of these strategies (when mentioned)

According to the authors of the studies [36]–[39], [41], the reported strategies helped to prevent HCWs and patients from being infected by the SARS-CoV-2 virus. The decision tree algorithm and the MeNTS scoring tool are considered as safe, efficient, valuable, and easy to implement tools to ration PPE, and to ensure optimal HCWs’ safety [38]. The two preoperative COVID-19 testing protocols were implemented rapidly and seem to have protected HCWs and patients from potential operative and perioperative COVID-19 exposures [36]. Self-collection of nasal and throat swabs were determined to be easy to perform and highly acceptable by the patients themselves, while providing patients with easier access to testing, increased the safety for both patients and staff by reducing mutual exposure and reduced the requirement for PPE [37].

#### 1.2. Space reorganization

##### Effects of the pandemic on hospital organizational routines

Since the beginning of the crisis, some hospitals have been quickly converted to COVID-19 care centers and some departments closed or shifted to assist patients with COVID-19 [32], [35]. This often resulted in frequent reorganization of physical space.

##### Strategies implemented to address these effects

- Creating dedicated high-risk COVID-19 areas. In Singapore and Italy, some hospitals partitioned their emergency departments (ED) into “higher risk” and “lower-risk” areas with dedicated staff and equipment and set up specific traffic flows for HCWs and patients [34], [43].
- Undertaking infrastructural modifications to accommodate the increasing number of COVID-19 cases. A study led in Singapore mentions the introduction of a respiratory surveillance ward (RSW) to facilitate the triage of patients presenting with respiratory syndromes [7]. An orthopedic center in Italy also quickly set up a dedicated COVID-19 operating room with reserved beds to isolate COVID-19 patients and provide HCWs with high-level PPE [32]. In Portugal, some emergency departments used alternative areas, such as tents or drive-in stations, for testing [44].

##### Impacts of these strategies (when mentioned)

These strategies have been judged as having positive impacts by limiting COVID-19 transmission within hospitals and protecting both patients and HCWs. In the Singapore General Hospital, since the division of the ED in high-risk “fever areas” and lower-risk zones, no cases of nosocomial transmission from intra-ED exposure were observed [43]. And the RSW has been successful in containing patients with COVID-19 in designated areas where appropriate PPE and infrastructural enhancements could reduce nosocomial transmission [7].

#### 1.3. Personal protective equipment and resources management

##### Effects of the pandemic on hospital organizational routines

The use of appropriate PPE by both professionals and patients is essential in minimizing nosocomial transmission of SARS-CoV-2 [7], [43], [45]. A single case of COVID-19 can result in the quarantine of large numbers of patients or HCWs, further straining hospital resources [7]. However, basic PPE was not always available in many medical institutions dealing with COVID-19 patients [8], [9], [33], [45]–[52]. These shortages can lead to the re-use or the inappropriate use of PPE by HCWs [33], [47], higher risk of infection and level of anxiety among HCWs [33], [51], [52], and physical injuries [46], [53].

##### Strategies implemented to address these effects

- Applying a risk-stratified approach to rationally conserve PPE and avoid infection. A large tertiary hospital in Singapore divided its ED in different zones: higher risk zones, or “fever areas”, in which all patients presenting to the ED with a respiratory syndrome or undifferentiated fever were classified and where HCWs had to use full PPE (i.e., N95 masks, eye protection or face shields, disposable gown and gloves); lower risk zones (e.g., triage areas, corridors of fever areas, observation ward, and critical care area) where ED staff wore N95 masks; and other low-risk areas where the usage of a surgical mask was made the mandatory minimum standard [43]. In California, the department of surgery of the Stanford University Medical Center developed an algorithm for PPE use by requiring a fitted N95 respirator mask in addition to droplet PPE (i.e., gown, gloves, eye protection) for the entire team managing emergency cases or COVID+ patients, and droplet PPE for all cleaning personnel [38].

##### Impacts of these strategies (when mentioned)

According to the authors, the “risk-stratified” approach has proven to ensure HCWs’ safety and a rational use of PPE in a resource-constrained setting. Managing all patients presenting with respiratory syndromes in designated fever areas with upgraded PPE and better bed spacing of patients prevented patient-to-staff transmission during the pandemic [43].

#### 1.4. E-health and the use of technologies

##### Effects of the pandemic on hospital organizational routines

Since the beginning of the pandemic, a radical increase in the use of e-health has been reported [30], [31], [40], [54], [55]. Telemedicine has proven to be an effective way to increase and expand patients’ access to care over a short period of time [55], decrease interpersonal contact [31], avoid overwhelming burdens on healthcare facilities [31], and provide care at times of shortages in human resources or equipment [55]. It has shown high rates of satisfaction among both patients and clinicians [30], [55], [56]. However, there are some limitations: it is still not widely available for all HCWs [9], professionals have not always been trained on how to perform effective teleconsultations [40], there are financial barriers such as the lack of reimbursement for most telemedical procedures [31], and patients must have access to an internet connection and be comfortable with the use of technologies [31], [57].

##### Strategies implemented to address these effects

- Implementing telephone follow-up and teleconsultation with electronic medical records. In India, a multi-tier ophthalmology hospital network set up a three-level triage protocol of teleconsultation calls with a dedicated telel⍰consult team of ophthalmologists, using the patient information retrieved from the electronic medical record (EMR) system [54]. In Poland, at the ambulatory clinic of implantable devices (ACID) of the Central Teaching Hospital, physicians phoned and interviewed patients about their health prior to planned visits and made further treatment decisions based on the interview and available EMR [31].
- Using a virtual care program to manage outpatients with COVID-19. In Toronto, the Sunnybrook Health Sciences Centre developed the COVID-19 Expansion to Outpatients (COVIDEO) program to provide ongoing care for outpatients diagnosed with COVID-19, and to follow and assess them via the Ontario Telemedicine Network virtual care platform or by telephone [57].

##### Impacts of these strategies (when mentioned)

Teleconsultations and access to patient information from EMR enabled a timely response, by handling a large volume of calls efficiently and ensuring continuity of care in the absence of physical visits to the hospital and information access to the patients [54]. The use of EMR has been judged as paramount given the importance of immediate access to information about the patient [31], [54]. In both studies, most patients gave positive feedback on teleconsultations. The COVIDEO program allowed HCWs to plan for a safe and controlled hospital transfer for those showing signs of clinical deterioration, but the authors identified several implementation challenges including increased number of patients, administrative challenges of booking virtual visits, and the fact that older people may not be as comfortable using technologies [57].

### 2. Hospital dimension: Human resources

#### 2.1. Reorganization of professionals’ work

##### Effects of the pandemic on hospital organizational routines

The COVID-19 pandemic has demanded increased focus on resource allocation and on the roles in which HCWs function [48]. Several physicians reported working outside of their normal scope of practice [48]. Heavy workloads impacted their work quality [8], induced stress and high risks of infection [8], [12], [48], [58], [59]. The contamination of HCWs resulted in a reduced workforce available for daily activities, coverage for on-call duties, and a general weakening of the response capacity [29], [60].

##### Strategies implemented to address these effects

- Redeploying HCWs and recruiting new workforce. A study led in different US urology departments documented how several specialists’ physicians were redeployed to “frontline COVID-19 services”, most commonly to ICUs and emergency departments [40]. Also, due to the drastic surge of patients worldwide, reinforcements strategies have been implemented in Europe, Asia and North America [8], [40], [59], [61]–[63]. Many countries involved students or physicians in training to join the workforce, largely regardless of their specialization [8], [62].
- Implementing specific measures to reorganize HCWs’ work. In Italy, the Operative Unit of Occupational Medicine of the University Hospital of Bari made changes in HCWs’ work to avoid overcrowding or infection risks, such as reducing the number of HCWs in rooms with suspected and/or confirmed COVID-19 cases, organizing necessary bedside medical and surgical procedures in advance, and performing nasopharyngeal swabs by a single HCW per work shift [41].

##### Impacts of these strategies (when mentioned)

According to the authors of the studies, these strategies helped to face the sudden surge of COVID-19 patients and to alleviate HCWs’ work. However, mandatory redeployments can impact HCWs’ motivation [40], and inexperience in such urgent situations may be particularly stressful for students graduating directly to responsibilities during the pandemic crisis [62]. Both can impact HCWs’ work quality.

#### 2.2. Knowledge and training of HCWs

##### Effects of the pandemic on hospital organizational routines

Some studies show the moderate-to-poor or the inadequate level of knowledge of HCWs about the COVID-19 pandemic or its management [47], [49]. Poor level of knowledge can be due to limited training [8], [14], [46], [64], or can be explained by misinformation due to the use of social media [47]. This can result in dissatisfaction among HCWs [46], higher levels of stress and anxiety [8], [14], and incorrect or irrelevant practices leading to higher risks of infection for HCWs [60]. HCWs want to be better prepared for emergencies [28], [49], and gain confidence in their role in reducing the risk of transmission [60].

##### Strategies implemented to address these effects

- Providing and facilitating online training for HCWs. Two studies, led in Denmark and in China, analyzed the differences between “traditional teaching” (i.e., instructor-led training) and video training in their effectiveness to handle training disruptions during the pandemic [65], [66]. In both strategies, the group of participants who watched training videos could freely watch again at home.

##### Impacts of these strategies (when mentioned)

The strategy enabled HCWs to access training, and information during the COVID-19 pandemic. The studies found no significant difference between the two types of training. The participants who received video training were on average as competent as those who received instructor-led training in-person. HCWs show high satisfaction with the use of video training [65] that has proven to be a resource-efficient way of reaching all relevant personnel without requiring face-to-face training [66], both in terms of time and money spent.

#### 2.3. Well-being and mental health of HCWs

##### Effects of the pandemic on hospitals’ professionals

Several studies found a high prevalence of sleep disorders, stress, anxiety, and depression among HCWs during the COVID-19 pandemic [12], [28], [33], [48], [52], [59], [61], [63], [67]–[72], for which students [8], [13], [56], [62], doctors [60], [70], non-frontline HCWs [73], and female HCWs [45], [63], [69], [74], [75] were greater risks to experience. Psychological distress and anxiety can be caused by: the fear of being infected and contaminating their families [8], [14], [29], [48], [58], [60], [64], [76]–[78]; the lack of knowledge and unavailability of clear protocols [28], [45], [58]; shortages of medical supplies and effective PPE [13], [14], [33], [40], [45], [52], [58], [63]; heavy workloads [13], [14], [28], [59], [60], [68], [78]; the disruption of training programs [14]; quarantine measures and isolation [56]; the overwhelming quantity of information in the media, including social media [62], [77]; concerns about their financial situation, mostly for surgeons because of the reduction of surgical and clinical activities [56]. Some studies reported measures considered to be effective in reducing HCWs’ anxiety: institutional support and recognition of their work by the medical profession and hospital management [58]; family or social support [13], [40], [58], [59], [62]; better virus prevention and transmission knowledge [58]; implementation of clear disease prevention guidelines [58]; a positive working environment (e.g., colleagues encouraging each other during work shifts, writing good-luck messages on personal protective equipment, enjoying lunches provided by social volunteers, communicating with others on social media…) [28], [33], [58].

##### Strategies implemented to address these effects

- Providing HCWs with institutional or family psychological support. A study from China reported the use of intervention teams to provide psychological services to HCWs [74]. Of all the participants of the study, 36.3% accessed psychological materials (e.g., brochures, books), 50.4% psychological resources available through media (e.g., online push messages on mental health self-help coping methods), and 17.5% participated in counseling or psychotherapy.
- Setting up a collaborative and helping crisis cell for HCWs in need. In France, a crisis cell was created for all residents of the French Association of Urologists in Training (AFUF) in need, to connect physicians in training with psychiatry residents 24 hours a day [8].
- Improving information dissemination. In China, the Second Affiliated Hospital of Guangxi Medical University provided access to various online platforms with medical advice to share information on how to decrease the transmission risks between patients [68].
- Enhancing HCWs’ personal resilience. An international study focusing on COVID-19 impacts on spine surgeons worldwide provided examples of coping strategies implemented by HCWs themselves, such as spending time with family, physical activity, reading, and practicing meditation or spiritual/religious activities [48].

##### Impacts of these strategies (when mentioned)

The studies demonstrated the negative effects of the COVID-19 pandemic on HCWs’ well-being and mental health without reporting the impacts of the strategies implemented.

### 3. Hospital dimension: Information and communication

#### Effects of the pandemic on hospital organizational routines

In moments of public health crisis, communication “is as crucial as medical intervention” [79]. The quantity of information sources and the rapid evolution of the pandemic has made it difficult to distinguish misinformation from valid information. This has resulted in enormous implications from delaying appropriate care to a disregard for evidence-based interventions [79]. Data shared online, particularly through social networks, can cause considerable information pollution [45], [47], [64], [79]. However, a study led in India demonstrated that social media could play an important role in information dissemination during emergencies and lead to concrete behavioral changes [79].

*No strategies or impacts related to information and communication practices have been found in the selected literature*.

### 4. Hospital dimension: Finance

#### Effects of the pandemic on hospital organizational routines

The COVID-19 crisis has put a massive burden on healthcare systems [77]. Since its beginning, there has been a shift of attention and resources from entire healthcare systems to focus solely on the COVID-19 response. However, this certainly implies opportunity costs of not solving health problems that should have been resolved during this period [44]. The cancellation or postponement of medical and surgical procedures leads to the accumulation of unmet or unresolved needs, to the rise of complexity and severity of some pathologies, and to increased health and economic risks.

*No strategies or impacts related to hospitals’ financing have been found in the selected literature*.

## DISCUSSION

### Main findings

#### Hospital dimensions and gaps in the literature

In the selected studies, almost all the attention was focused on the two following hospital dimensions: 1) “human resources”, including HCWs’ well-being and mental health, work reorganization, and HCWs’ knowledge and training; and 2) “planning, management and security”, including protection protocols for patients and professionals’ safety, space reorganization, PPE and resources management, and e-health and the use of technologies. The “information and communication”, “finance” and “governance” dimensions were mentioned in a small number of studies and often lacked rigorous analysis. Some studies also focused on HCWs’ professional values (i.e., values that are particularly important for HCWs and encourage them to work), part of the health systems’ “software” dimension [80]. These studies, mainly conducted in China, focused on the Chinese workforce, and referred to specific vocabulary and expectations. Chinese HCWs are expected to “exude a sense of responsibility and collective action” in the fight against the epidemic [68]. Their “willingness” to practice during the crisis is even higher than in previous outbreaks [61], their “social and moral responsibilities and professional obligations” being the most important factors that motivate them to continue working during the COVID-19 outbreak [58].

Most of the studies were directly written by HCWs working in different hospital settings during the pandemic. Participation by HCWs in the process of knowledge creation can be an invaluable tool, by enabling the creation of a “collective space for health professionals to reflect on and improve their practices” [81]. However, this process could also represent a scientific bias that can bring into question the neutrality of the scientific research process.

There were limitations in the selected studies. Firstly, we found that only one study used a conceptual framework to further analyze its data. Research is not sufficiently supported by theories and analytical frameworks [82], even though their *a priori* or *a posteriori* use could allow researchers to comprehensively assess the complex and dynamic process of healthcare access [83], [84]. Secondly, inequalities were ignored. The term “inequalities” was only used once in the selected studies, reflecting the more global lack of consideration of inequalities in the design of public health interventions during a crisis period [85]. Almost all the articles about hospitals’ professionals were focused on nurses, doctors, or specialist practitioners (e.g., surgeons), neglecting other professions such as paramedics, caregivers, or administrative professionals. Similarly, gender differences were rarely discussed in the selected studies. It suggests a “gender blindness”, the systemic failure to acknowledge gender differences in health [86], [87].

#### Hospitals’ resilience outcomes and processes

The selected studies were more descriptive in nature and all the reported impacts of the strategies were positive. We found more observations than specific strategies evaluated, with only a few studies performing any kind of systematic analysis to assess the impacts of these strategies. In the COVID-19 context, the rapidly evolving situation made it difficult to evaluate the impacts of recently implemented strategies or interventions. The goal of most of the included studies was to share knowledge as quickly as possible, but the lack of rigorous analyses is problematic [88] and does not allow the identification of which strategies are effective.

Few studies referred to actions taken in anticipation of the pandemic (strategies/impacts, before any effects) with most having reported cases of “inaction” (effects without any strategies). Most of the articles highlighted how some processes undertaken during the pandemic led to absorptive processes, healthcare facilities trying to absorb the shock by implementing short-term measures. For example, the creation of a crisis cell for French urologist residents or the use of specific algorithms (e.g., algorithms to facilitate patients’ prioritization or to rationally use PPE), in different countries, seem to be temporary measures that may not lead to global transformative processes. However, several articles attempted to increase healthcare access in ways that could potentially lead to a positive transformation process. For example, articles focusing on e-health mentioned that the current experience of tele-medicine, with the implementation of tele-consultations and the use of electronic medical records, could provide valuable insights to the possibility of managing patient follow-up visits remotely in the future [54]. Further research is needed to examine whether these resilience processes have led to improved access to healthcare in hospitals following the pandemic.

### Implications for professionals

The authors of the selected studies gave recommendations for practice to HCWs and hospitals’ leaders. An overview of these recommendations is depicted in (Table S2: Recommendations). We found more recommendations than specific strategies in the studies, which need to be taken with caution given the lack of evaluation in terms of their effectiveness.

During the COVID-19 pandemic management, obstacles encountered by hospitals and their personnel may or may not have been overcome for various reasons. The current pandemic provides important opportunities to propose high-quality lessons [89] learned from both positive and negative experiences. This work is essential to guide professionals’ practices, especially with the arrival of future waves of the pandemic.

### Implications for research

This scoping review highlights the need for more rigorous intervention research and evaluation, and the inclusion of multi-disciplinary teams involving social science researchers and epidemiologists [90]. A structured research agenda to inform health policy and system responses to COVID-19 should include hospital resilience research [91]. The sharing of high-quality lessons learned is crucial to designing appropriate policies to contain the COVID-19 pandemic.

### Limitations of the study

Firstly, as we chose to conduct a scoping review, the simplification of some steps of the systematic review to speed up the process can influence its rigor. To make these risks explicit and allow transparency, we gave a very detailed description of the method. Secondly, because of the very large amount of data available, we decided to exclude grey literature and preprints from our searches, therefore we could also have missed pertinent studies. Thirdly, we faced the analytical challenge of causality, for epistemological and methodological reasons. The first reason is the use, essentially, of a qualitative approach, and the second reason is the pandemic context, which did not facilitate a longitudinal data collection.

## CONCLUSION

There is a wide range of studies related to hospitals’ resilience that help us to understand which strategies have been implemented to face the COVID-19 pandemic. Empirical scientific literature can provide important lessons on protection protocols, space reorganization, PPE and resources management, e-health and the use of technology, work organization, training, HCWs’ well-being and mental health, information and communication, governance, and finance. However, data must be updated regularly to report the evolutions of the strategies implemented, and to provide health decision makers and professionals with recent and relevant lessons learned from concrete hospitals’ experiences to help them to better tackle new COVID-19 waves or future outbreaks. Further research on healthcare access is also needed to study the global resilience of hospitals.

## Supporting information

Supplementary Table 1: Description of selected studies

Supplementary Table 2: Recommendations

## Data Availability

Data referred to in the manuscript are derived from public domain resources.

## Acknowledgements

We would like to thank France Nadeau and Julie Desnoyers, librarians from the University of Montreal, for their availability and their help in writing the research strategy, and Flore-Apolline Roy who designed the world map.

